# Multi-site Validation of a SARS-CoV-2 IgG/IgM Rapid Antibody Detection Kit

**DOI:** 10.1101/2020.05.25.20112227

**Authors:** Christopher Minteer, Arnau Casanovas-Massana, Tao Li, David McDonald, Linda Wang, Si Hui Pan, David Caianiello, Jesse Collinski, Edward deRamon, Robert Hale, Rebecca Howell, Jason Ray, Joseph Vinetz, Morgan Levine, Albert I. Ko, David A. Spiegel

## Abstract

Deaths from coronavirus disease (COVID-19) have exceeded 300,000 persons globally, calling for rapid development of mobile diagnostics that can assay widespread prevalence and infection rates. Data provided in this study supports the utility of a newly-designed lateral flow immunoassay (LFA) for detecting SARS-CoV-2 IgM and IgG antibodies. We employed a clinical cohort of 1,892 SARS-CoV-2 patients and controls, including individuals diagnosed by RT-qPCR at Yale New Haven Hospital, The First Affiliated Hospital of Anhui Medical University, the Chinese Center for Disease Control and Prevention of Hefei City (Hefei CDC), Anhui Province (Anhui Province CDC), and Fuyang City (Fuyang CDC). The LFA studied here detects SARS-CoV-2 IgM and IgG antibodies with a specificity of 97.9–100% for IgM, 99.7–100% for IgG, and sensitivities ranging from 94.1–100% for patients >14-days post symptom onset. Sensitivity decreases in patients <14-days post symptom onset, which is likely due to lower IgG/IgM antibody levels in this population. Finally, we developed a visual intensity reporting system that we believe will be suitable for laboratory and point-of-care settings, and will provide granular information about antibody levels. Overall our results support the widespread utility of this and other LFAs in assessing population-level epidemiological statistics.

## Background

Coronavirus disease 2019 (COVID-19) has now spread to nearly every nation in the world [1]. As of May 22^nd^, 2020, more than 5 million cases have been reported globally, with more than one and a half million cases in the United States alone. Worldwide deaths from COVID-19 have reached 300,000 persons, although true prevalence and mortality is likely much higher, as limited diagnostic test implementation and the prevalence of asymptomatic carriers has masked the virus’s full impact [2].

SARS-CoV-2, the virus that causes COVID-19, is a respiratory pathogen that is readily transmitted from droplets and direct contact [1,2]. The disease course of COVID-19 is highly variable. Many patients have relatively mild symptoms, while certain high-risk groups, such as the elderly and immunocompromised, can develop multi-organ system complications that often results in death. For example, the CDC reports age-related mortality rates ranging from 10-27% for persons aged >85 years, 3-11 *%* for persons aged 65-84 years, and <1% among persons aged <54 years [3].

Efforts by the United States government to slow the spread of the virus have relied largely on social distancing. Despite these efforts, nearly 25,000 new infections occur each day in the United States as of May 22^nd^, 2020. Furthermore, quarantine measures decrease quality of life and have negative economic impacts. Widespread access to diagnostic tests will be critical for restoring social normality and mitigating the harmful impact of the virus. In particular, diagnostic tests for immunological status, which measure IgG and IgM antibody responses, can provide essential information about an individual’s past exposure to SARS-CoV-2 while also assessing the risk for subsequent infection. Most currently available antibody tests require complex laboratory settings; yet because 80% of all infected individuals may be asymptomatic, a reliable and easy-to-use diagnostic assay is desirable for deployment among the general population [2].

Immunochromatographic lateral flow assays (LFAs) have the potential to serve as such a tool [4-6]. LFAs can be readily conducted by untrained individuals, only requiring simple fingerstick blood draws, to generate an easy-to-interpret readout (Figure 1) in no more than 30 minutes. The LFA studied here provides two separate outputs – for both IgG and IgM – and therefore provides nuanced data related to current and past infections, while also helping to assess potential immunity [5,7]. SARS-CoV-2 specific IgM antibodies predominate immediately after infection while IgG antibodies are produced in the later stages, approximately two weeks post initial infection [1,2,4]. Although it is currently unclear whether antibody responses alone provide immunity to SARS-CoV-2 reinfection [8,9], neutralizing antibodies have been observed in several cases, and antibody levels are also likely to serve as an index of other protective immune responses [7,10]. After the United States Department of Health and Human Services began issuing emergency use authorizations (EUA) for immunoassay tests for SARS-CoV-2 diagnostics, many manufacturers have submitted applications seeking approval. However, according to recent studies, comparative performance of these tests is variable [5].

**Figure 1:**
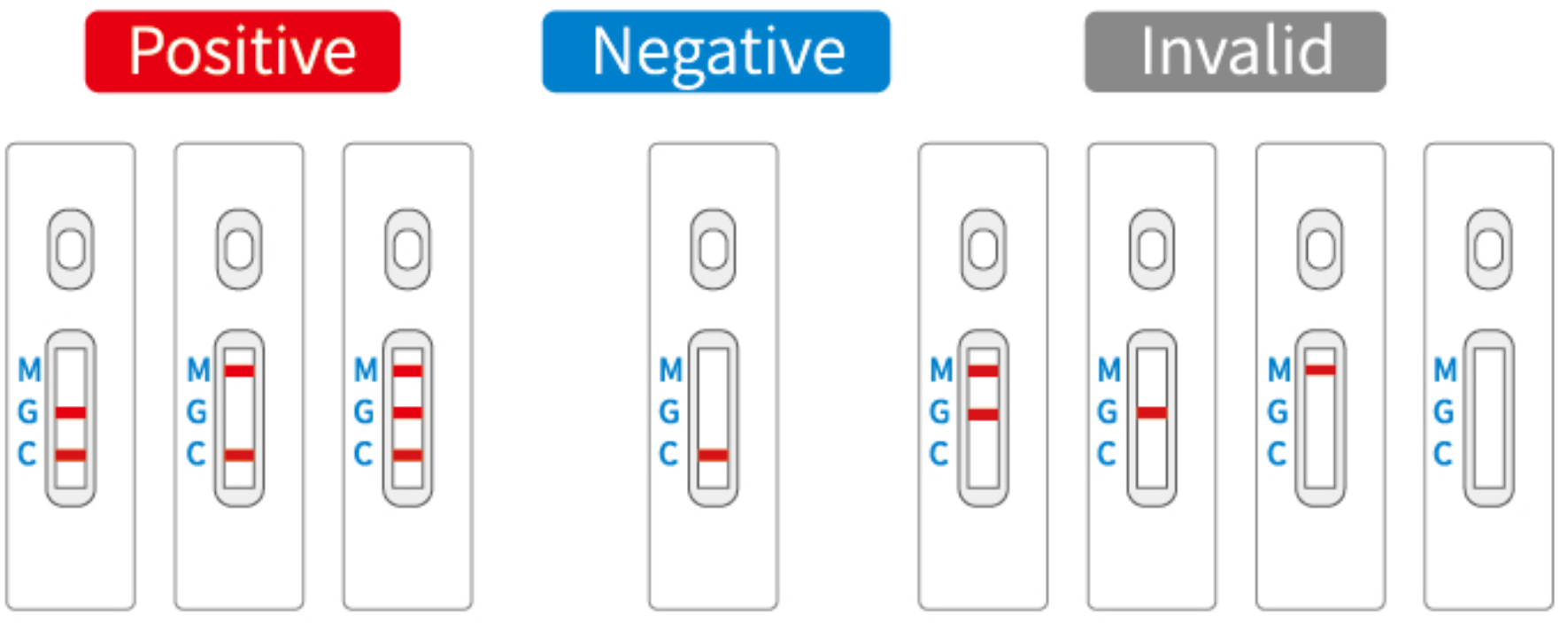
SARS-CoV-2 IgM & IgG colloidal gold immunochromatography lateral flow assay

Here we describe experiments to validate the lateral-flow-based immunoassay (LFA) for IgM and IgG SARS-CoV-2 antibodies, manufactured by Biohit Healthcare (Hefei) Co., Ltd. (subsequently referred to as Biohit). We chose this assay for study because of its strong performance in pilot studies. The sample population at Yale was well distributed by age, gender and infection time, which is defined as the number of days between patient-reported symptom onset and sample acquisition (Figure 2). Antibody titer in this cohort was found to correlate with infection time, indicating that the study population is representative of the general population (Figure 3). Furthermore, patients in the four study cohorts evaluated in China comprise a large number of diverse clinical presentations; in addition to patients with SARS-CoV-2, patients suffering from non-SARS-CoV-2 respiratory illnesses, pregnant females, and healthy controls were also tested.

**Figure 2:**
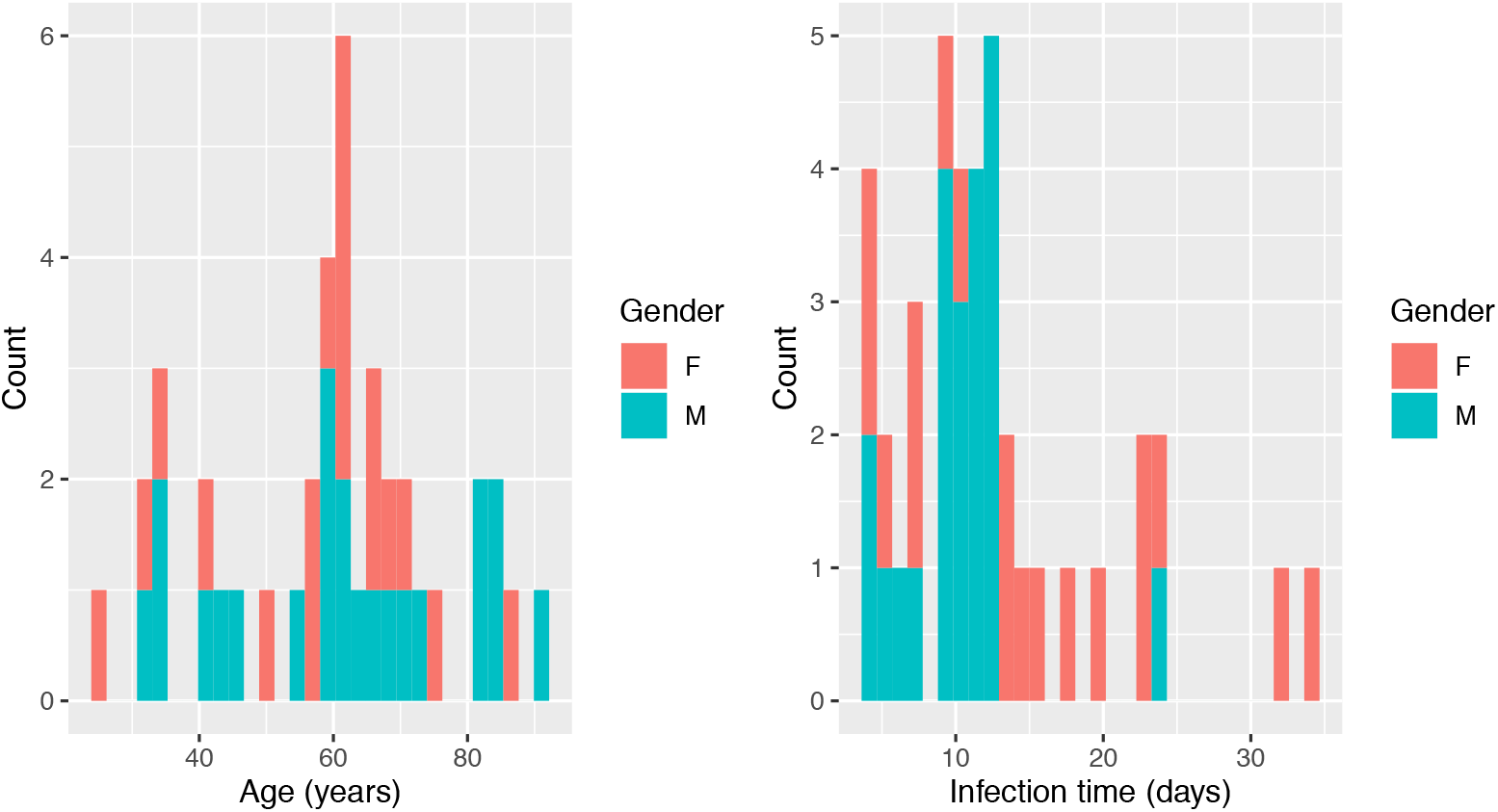
Clinical characterization of SARS-CoV-2 positive patients from Yale New Haven Hospital. Age range: 26-92 years old and Mean age: 59.6 years old, Infection range: 4-34 days and Mean infection time: 12.4 days

**Figure 3:**
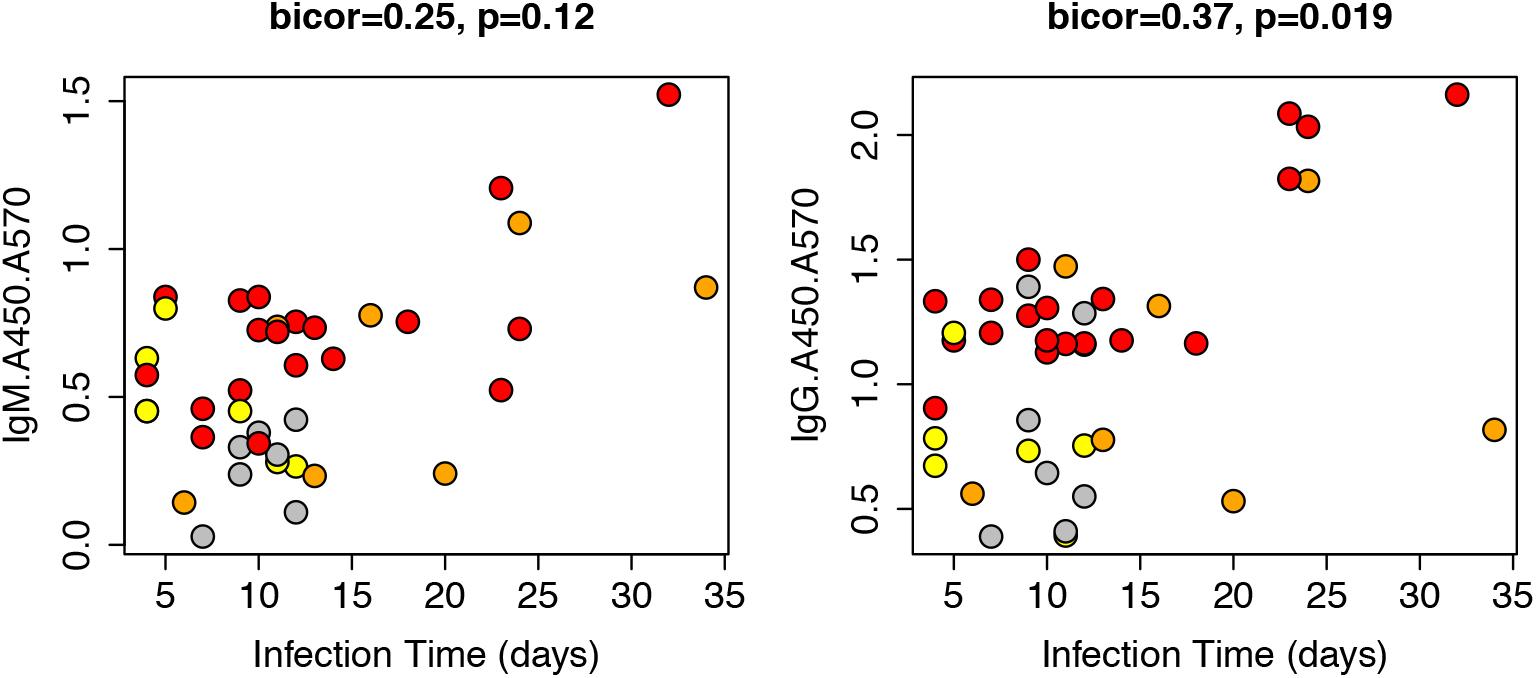
IgM and IgG titer for SARS-CoV-2 positive samples plotted as a function of infection time for Yale New Haven Hospital samples. Gray = False negative (Visual intensity score = 0), Yellow = Positive (Visual intensity score = 1), Orange = Positive (Visual intensity score = 2), and Red = Positive (Visual intensity score = 3)

The LFA under study is highly sensitive and specific for both IgG and IgM antibodies recognizing either the S1 domain of the spike protein (containing the RBD) or the nucleocapsid protein (Figure 4). Sensitivity is lower at earlier infection times and correlates with decreased antibody titers as measured by ELISA. In other words, false negatives occur with greater frequency in samples with low antibody titers. These results are consistent with observations that IgM levels are higher at earlier times during the disease course whereas IgG levels increase at later disease stages [4].

**Figure 4:**
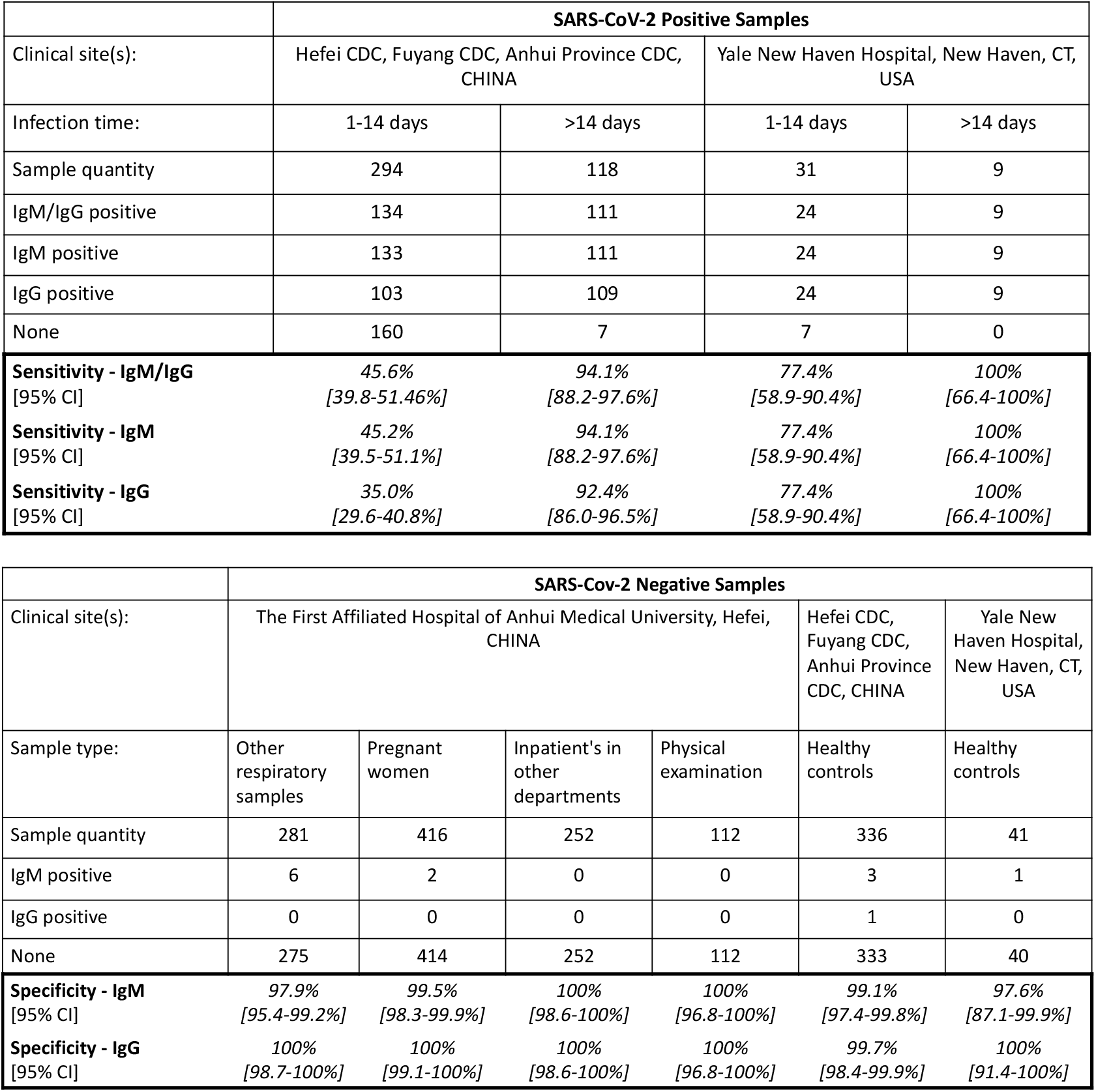
Sensitivity and specificity of SARS-CoV-2 patients and controls. IgM and IgG detection were determined using LFA. **2 samples did not indicate infection time from Yale New Haven Hospital SARS-CoV-2 patient records, thus results were omitted.

We also developed a visual intensity score (VIS, Figure 5), which correlates with ELISA titer (Figure 6). This correlation supports that LFA band intensity is measuring antibody levels in serum. We develop an automated intensity score (AIS), which involves computer-assisted analysis of LFA result photographs taken on a smartphone. The AIS correlates well with the VIS, and is particularly effective for darker intensity LFA bands (Figure 7). The VIS tends to fail, however, in distinguishing low intensity positives from negatives, suggesting that visual inspection is superior to computer-assisted assay analysis. This finding may have implications for how best to implement LFA assays in point-of-care and home settings, and may also suggest a role for remote healthcare professionals test result analysis via telemedicine.

**Figure 5:**
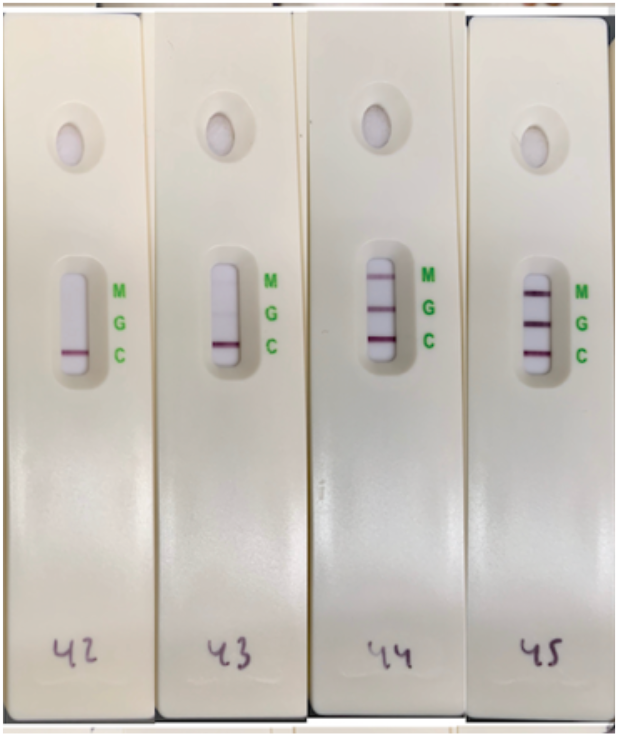
Representative photo of SARS-CoV-2 LFA results. Note, Sample 42= Negative (0), Sample 43= Weak Positive (1), Sample 44= Positive (2), Sample 45= Strong Positive (3)

**Figure 6:**
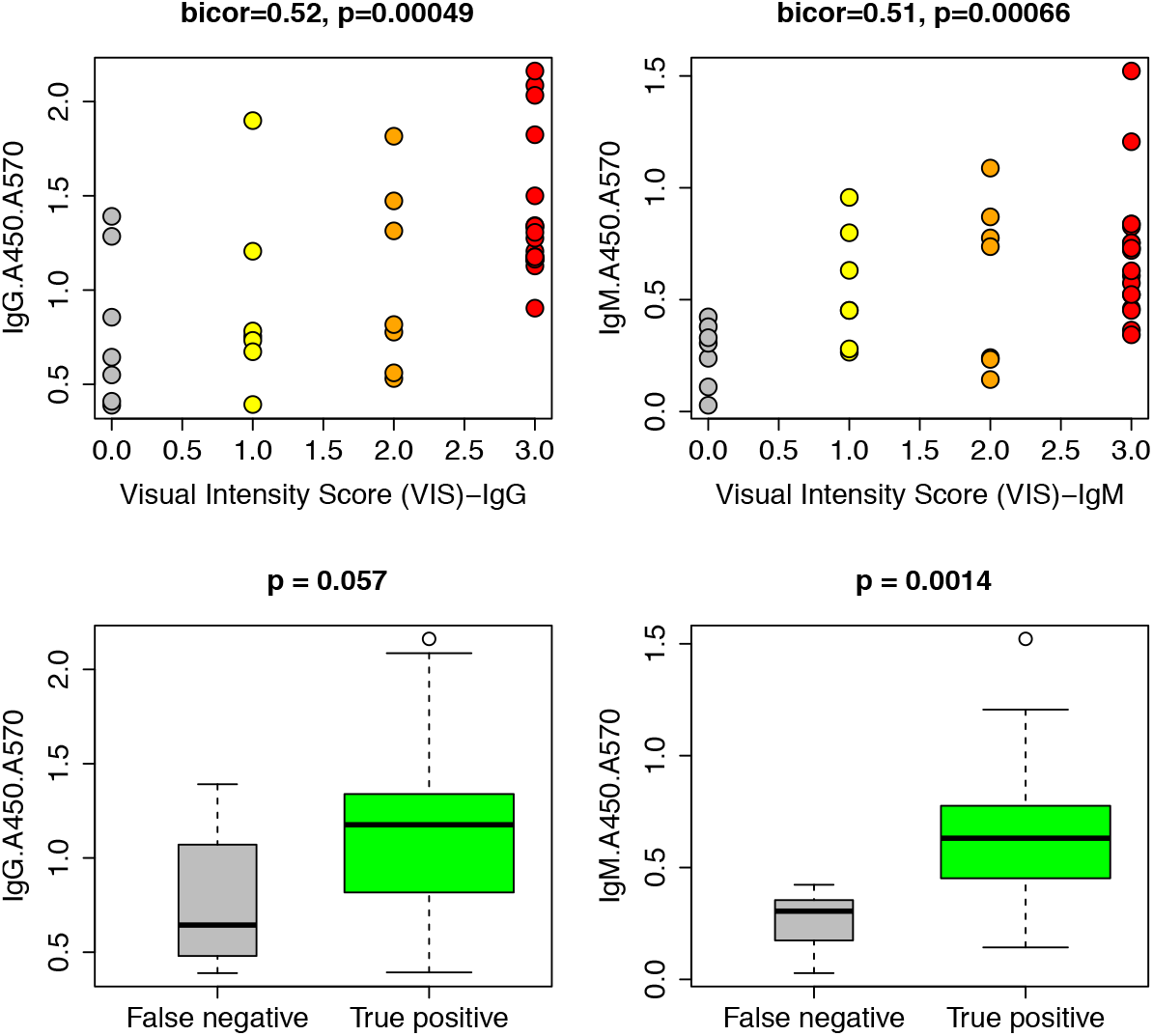
Visual intensity score and concordance compared with ELISA IgM and IgG titer of SARS-CoV-2 positive samples. Gray = False negative (Visual intensity score = 0), Yellow = Positive (Visual intensity score = 1), Orange = Positive (Visual intensity score = 2), and Red = positive (Visual intensity score = 3)

**Figure 7:**
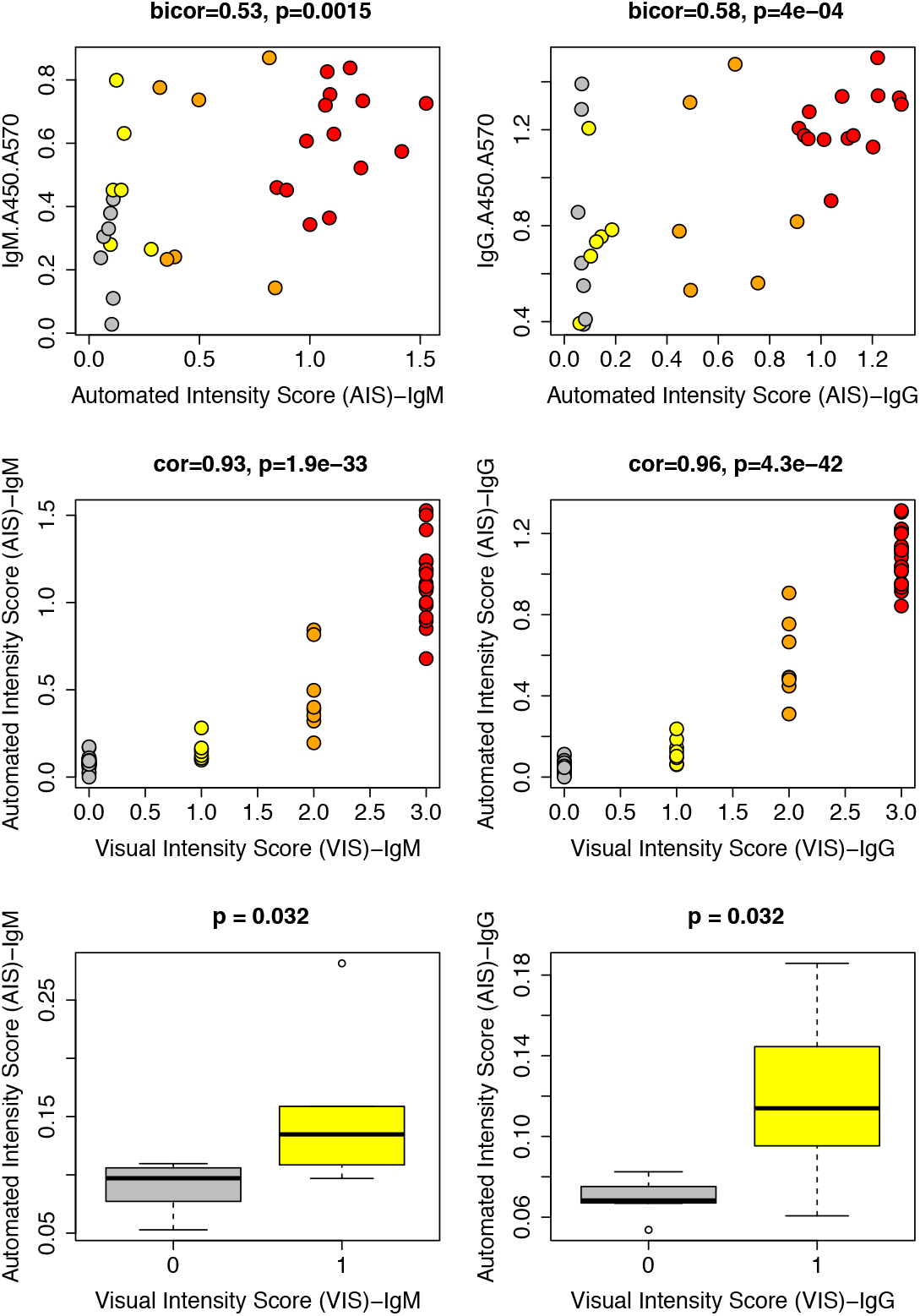
IgM and IgG band quantification for SARS-CoV-2 positive patients compared to ELISA titer and visual intensity score. Gray = False negative (Visual intensity score = 0), Yellow = Positive (Visual intensity score = 1), Orange = Positive (Visual intensity score = 2), and Red = positive (Visual intensity score = 3)

We then conducted control experiments to confirm test specificity, robustness and matrix choice. Comparison between whole blood and serum samples for twelve separate patients revealed perfect correlation, confirming serum and whole blood are appropriate matrices for application in this LFA (Supplemental Figure 2). Time-course experiments indicate that positive bands begin to appear as early as 30 seconds, and remain stable by VIS after 15 minutes (Supplemental Figure 3). LFA bands remain visible for both IgG and IgM at concentrations up to four times the recommended loading (40 μL versus 10 μL) and down to a 1:100 dilution (Supplemental Figure 4). We also demonstrate isotype specificity of the LFA through Protein A/G depletion experiments. These studies are consistent with the conclusion that the IgG and IgM band readouts on the LFA test measure of IgG and IgM levels respectively (Supplemental Figure 5). Finally, we tested a large panel of interfering substances and none were found to alter test performance (Supplemental Figure 7).

Taken together, these studies support the effectiveness of the LFA under study.

## Material and Methods

### 1.1 Test development

#### Colloidal gold immunochromatography lateral flow assay

SARS-CoV-2 IgM & IgG antibodies were detected using colloidal gold immunochromatography lateral flow assay technology, consisting of sample pad, conjugate release pad, nitrocellulose membrane with immobilized test line 1 (IgM), test line 2 (IgG) and control line, and finally absorbent pad.

Both SARS-CoV-2 IgM & IgG antibodies were detected simultaneously using SARS-CoV-2 recombinant antigens and mouse anti-human IgM & IgG antibodies. Specifically, colloidal gold-bound SARS-CoV-2 recombinant antigen (Nucleocapsid Protein and Spike protein (RBD)) reacts with SARS-CoV-2 IgM and/or IgG at the conjugate release pad, then the complex is chromatographed along the nitrocellulose membrane as it reaches each test and control line. Note, the control line detects mouse anti-human IgM and IgG from the conjugate pad and must be present to indicate a functioning test.

A positive result occurs when the colloidal gold SARS-CoV-2 recombinant antigen-antibody complex is bound to the IgM or IgG detection test line(s), producing purple-red band(s). A negative result occurs when the colloidal gold SARS-CoV-2 recombinant antigen-antibody complex fails to form or is not present, such that both the IgM and IgG test lines are not visible (Figure 1).

### 1.2 Clinical population and research goal

#### Study approval

The 83 clinical samples from Yale New Haven Hospital were collected in accordance to the HIC-approved protocol #2000027690. Written informed consent was obtained from all participating patients and healthy controls. Permission was obtained from health and local authorities after they received an explanation of the purpose and procedures of the study. Written informed consent was obtained before participation and blood sampling from all adults and the parents of all participating children less than 18 years. Along with their parents’ consent, children older than 7 years provided written informed consent.

Samples from the Chinese cohorts were obtained from four sites: the First Affiliated Hospital of Anhui Medical University, the Chinese Centers for Disease Control and Prevention in Hefei City (Hefei CDC), Anhui Province (Anhui Province CDC), and Fuyang City (Fuyang CDC). Samples were collected at the First Affiliated Hospital of Anhui Medical University under ethical guidance from the hospital’s IRB (Quick-PJ 2020-04-18). Each of the three CDC sites served as a central repository for samples received from local hospitals all of which were collected in accordance with policies outlined by the National Health Commission of the People’s Republic of China [11]. Specifically, venous blood samples were collected from any patient displaying symptoms of the novel coronavirus. Those samples were then sent to the governing regional branch of the Chinese Centre for Disease Control and Prevention (CDC), including the three CDC sites participating in this study (Hefei, Fuyang, Anhui). All identifying patient data were removed before data was collected by researchers at Biohit.

Sample and site distribution are summarized in Supplemental Figure 1.

#### Clinical center background (Patient Selection)

Yale School of Public Health served as an independent validation site from the test manufacturer (Biohit healthcare (Hefei), Figure 2). Briefly, a total of 83 patients were enrolled: 42 patients who were positive for SARS-CoV-2 infection, as determined by RT-qPCR (age range 26–92y and mean age: 59.6y), and 41 patients who were negative by RT-qPCR and ELISA (age range 23–62y and mean age: 36.0y). SARS-CoV-2 patients were further selected on the basis of positive ELISA results such that ELISA values could serve as the “gold standard” in comparison with LFA results. Antibody titer in this cohort was found to correlate with infection time (Figure 3). These results are consistent with observations that IgM levels are higher at earlier times during the disease course whereas IgG levels increase at later disease stages [4], and indicate that the study population is representative of the general population. Again, sample and site distribution are summarized in Supplemental Figure 1.

Infection time is defined as the number of days from patient-reported symptom onset to the date of sample collection. Infection time at assessment ranged from 4 to 34 days, with a mean of 12.4 days, and comprised both acute and recovery responses. Because patient-reported symptom onset was not collected from 2 patients, we omitted these patients from all analyses involving the infection time variable.

A second cohort of 1809 samples was obtained from the First Affiliated Hospital of Anhui Medical University, Hefei CDC, Fuyang CDC and Anhui Province CDC. These patients included 412 patients who tested positive for SARS-CoV-2 and 1397 patients who tested negative for SARS-CoV-2 by RT-qPCR. Negative samples were further subdivided into 281 patients with non-SARS-CoV-2 respiratory infections (including Mycoplasma pneumoniae, parainfluenza virus, adenovirus, and influenza B virus; Age range: 2–99y and Median age: 51y), 252 patients with non-respiratory infections (including 30 cases of rheumatic immune system diseases and 20 cases of severe liver disease; Age range: 1–90y and Median age: 50y), 416 pregnant patients (Age range: 18–34y and Median age: 27y), 112 healthy patients who were undergoing routine physical examinations (Age range: 23–72y and Median age: 50y) and 336 healthy control patients with age not reported. Information about patient diagnoses and clinical sites is reported in Supplemental Figure 1.

At all sites, studies were run in a randomized, blinded fashion such that experimenters were unaware of all identifying patient information, including infection status.

### 1.3 Specimen collection and scope

#### Blood sampling and clinical diagnosis

Serum was the main matrix employed throughout this study, although whole blood from certain patients was used as a control. Venous whole blood was collected from study participants, and serum was obtained as follows: samples were acclimated to room temperature and centrifuged at 3500 rpm for 5 minutes. Following centrifugation, serum was collected and stored at ™20°C for antibody testing by ELISA or lateral flow test. SARS-CoV-2 diagnosis for admitted hospital patients at Yale New Haven Hospital was made by RT-qPCR on nasopharyngeal swabs collected upon hospital admission (Yale New Haven Hospital EUA approved method). Health care workers were tested for the presence of the virus in nasopharyngeal swabs and/or saliva by RT-qPCR using the 2019-nCoV_N1 and N2 primer sets [12] as detailed previously [13]. Chinese clinical sites used similar Fluorescence PCR methods for clinical diagnosis.

### 1.4 Test validation studies

#### SARS-CoV-2 s1 spike protein IgM & IgG serological ELISA quantification

ELISA assays for IgG and IgM antibodies towards SARS-CoV-2 were performed on patient plasma as described by Amanat *et al*. [14]. The capture antigen employed was the SARS-CoV-2 spike glycoprotein subunit 1 (S1, Acro Biosystems #S1NC52H3). Detection antibodies employed included horseradish peroxidase (HRP)-conjugated anti-Human IgG (GenScript, #A00166) and horseradish peroxidase (HRP)-conjugated anti-Human IgM (Sigma-Aldrich, #A6907). Screening of plasma samples was performed with a 1:50 dilution. The cut-off was set with a confidence level of 99%, as described by Frey et al [15].

#### IgG knockdown

To confirm isotype specificity in the lateral flow immunoassay, IgG depletion studies were performed. Specifically, serum from a known IgG/IgM-positive patient was depleted of IgG by passage over a Protein G column, then again over a Protein A column. The eluent was then tested in the SARS-CoV-2 antibody LFA as described in Section 1.5.

#### Dilution experiments and time-course assays

Dilution series and time-course experiments (0–45 minutes) were used to determine the robustness and versatility of SARS-CoV-2 antibody detection. Briefly, serum was diluted in elution buffer by the factors indicated, then utilized as described below in Section 1.5.

### 1.5 Testing protocol

#### Instruction for lateral flow testing

The operators conducting the lateral flow tests were scientists with technical expertise and were blinded to all identifying information. Briefly, 10 μL of either serum or whole blood was pipetted onto the sample pad, followed immediately by the addition of 80uL of aqueous diluent (50 mM PBS, 10 g/L Casein, 0.05% Tween20, Proclin 300 0.3%, pH 7.4) by pipette to the sample. Lateral flow cassettes were immobilized on a lab bench and tests were conducted at room temperature (20°C).

#### Visual interpretation score (VIS) determination

Unless otherwise noted, the operator photographed the tests and determined the diagnostic outcome as either positive or negative for IgM and IgG after approximately 15 minutes. The operator also provided a visual interpretation score (VIS) on a scale of 0, 1, 2 and 3, with 0 being no intensity, 1 as weak intensity, 2 as medium intensity, and 3 as strong intensity.

#### Quantitative automated interpretation score (AIS) determination

Following initial test interpretation as described above, photographs were taken using an iPhone X smartphone. Results were then quantified in a blinded fashion by a different scientist, blinded to all identifying patient information, using LI-COR Image Studio. Briefly pixel intensities for IgG, IgM and control bands were quantified and data are reported as a ratio of IgG and IgM bands with respect to control as indicated.

### 1.6 Statistical methods and analysis

R was used for statistical analysis. When determining statistical significance, either the Kruskal Wallis test, which is a non-parametric multi group comparison test, or the Fisher test for two groups, were used. Confidence intervals (95%) for sensitivity and specificity were calculated as "exact" Clopper-Pearson confidence intervals.

## Results

### Test performance

#### 1.1 Sensitivity and Specificity

The performance of the SARS-CoV-2 LFA was evaluated using 1,892 total samples. Overall, the test achieved between 97.6–100% IgM specificity and between 99.7–100% IgG specificity (Figure 4). The sensitivity for IgG and IgM was found to depend on infection time. Consistent with previous reports [4], our study reports between 45.6%–77.4% sensitivity for short infection times (1-14 days) and between 94.1%–100% sensitivity for longer infection times (>14 days). Our data suggests that the decreased sensitivity observed in patients with <14-day infection times resulted largely from low antibody levels in these patients (Figure 3).

Control experiments explored differences between serum and whole blood, as well as time- and concentration-dependence. No changes were observed in test results comparing whole blood to serum (Supplemental Figure 2). Time course experiments showed that bands were visible as soon as 30 seconds after adding sample, but remained consistent between 10 and 45 minutes. For the majority of studies, we evaluated tests at approximately 15 minutes (Supplemental Figure 3). Dilution experiments (Supplemental Figure 4), performed for two separate patients, indicated that sample could be diluted up to 100-fold before IgM and IgG bands were found to disappear by visual inspection.

### Determining the utility of LFAs for SARS-CoV-2 antibody detection

#### 2.1 Visual interpretation of LFA testing results

Evaluation of test results were made by trained laboratory scientists who were blinded to all identifying patient information. Because LFAs are readily interpreted via visual inspection, we developed a semi-quantitative visual intensity score (VIS) wherein: 0 indicates a negative, 1 indicates a weak positive, 2 indicates a medium-intensity positive, and 3 indicates a strong positive (Figure 5). Indeed, the VIS for both IgG and IgM bands were found to correlate strongly with IgM and IgG titer, as determined by ELISA (Figure 6). To confirm isotype specificity for IgM and IgG, we conducted IgG depletion experiments using Protein G/A chromatography (Supplemental Figure 5). These studies indicated that depletion of IgG from SARS-CoV-2 positive samples resulted in abrogation of IgG band, consistent with the indicated isotype specificity.

#### 2.2 Test design and human factor implications

The utility of this test for at-home use will rely in part on its ease of interpretation. We employed LI-COR image studio to quantify the intensity of IgM and IgG bands from smartphone images. To this end, we developed an automated intensity score (AIS), as described in Materials and Methods. The AIS was found to correlate with ELISA IgM and IgG titer, as well as overall VIS (Figure 7). While the AIS performed well for strongly positive results, it had difficulties distinguishing weakly positive and negative signals (Figure 7). This distinction was better for IgG than IgM, but overall, our data suggests that automated mobile detection through a smartphone application could have difficulties differentiating weakly positive patients from true negatives, potentially increasing false negative rate. Finally, we considered confounding clinical variables that could alter LFA results.

### Clinical characteristics and potential confounders

#### 3.1 Specificity and endogenous interfering substances

A chief concern for implementing any diagnostic is understanding the risk and likelihood of false positives and false negatives. In the case of the SARS-CoV-2 LFA test, false positive results are particularly concerning because they can be incorrectly interpreted to indicate that a user is protected from subsequent infection. Therefore, it is imperative that the specificity is particular to SARS-CoV-2 and not other forms of respiratory infections. Overall, our results indicated an extremely low false-positive rate and therefore an excellent level of test specificity. For example, the test performed with 100% specificity for both IgM and IgG in patients with non-SARS-CoV-2 illnesses, and in healthy patients attending physical examinations (Figure 4). For patients with non-SARS-CoV-2 respiratory infections the test performed with a 99.5% specificity for IgM and 100% specificity for IgG. For pregnant women, the tests performed with 97.9% specificity for IgM and 100% specificity for IgG (Figure 4). Taken together, our data suggests that IgG is a more robust clinical endpoint for SARS-CoV-2 detection, whereas IgM from other infections or patient background may introduce specificity concerns.

We then analyzed various outcomes by ICU and mechanical ventilation status and found they did not correlate with LFA results, including VIS and AIS scores, or ELISA titer (Supplemental Figure 6). Finally, the LFA was evaluated for endogenous reactivity for common substances that SARS-CoV-2 patients may have been exposed to during treatment or prior to infection (Supplemental Figure 7). We report that the LFA test achieved 100% specificity for both IgM and IgG in whole blood, serum and diluent buffer samples spiked with mucin, blood based, antiviral, antibiotic or allergic system relief drugs and substances at varying pharmacologically relevant concentrations (Supplemental Figure 7). All in all, our data suggests that the studied LFA is highly specific to SARS-CoV-2 IgM and IgG antibodies, with robust sensitivity depending on infection time.

## Discussion

The decisions by government and public health officials to reopen global economies post COVID-19 will depend not just on active infections, but also on estimates of herd immunity acquired through prior infection. Understanding precise patterns of community spread of this infection will enable targeted public health interventions. Quantifying antibody exposures will help inform state and county prevalence rates, which provides authorities and employers with the ability to re-commence work and daily life. Quantifying exposure has been challenging to date due largely to the limited availability and prohibitive cost of high complexity diagnostics.

In just the past few months, multiple studies have clarified the role of antibodies, chiefly IgM and IgG, in the immune response to SARS-CoV-2 [4,5]. Importantly, IgM antibodies are readily produced as early as 1 day after infection, with IgG antibodies following at approximately 6 days. IgG responses increase exponentially between 12–15 days, at which point they begin to plateau; IgM responses plateau around 9–12 days and then subsequently decrease [4].

Our results suggest that LFAs, such as the one studied here, can accurately differentiate SARS-CoV-2 patients with nearly perfect specificity and high sensitivity (Figure 3 and 4). The observation that sensitivity is greatest for patients with longer infection times is largely explained by higher levels of antibodies found in these patients. This trend is consistent with recently published studies showing high variability in antibody levels at short infection times [4,16].

We also note a substantial difference in sensitivity between data collected at Yale versus at the Chinese sites (77.4% versus 45.6%) at early infection times (1-14 days). We hypothesize that this disparity results from the different “gold standard” assays employed in the two studies. All samples from the Yale site were pre-selected for ELISA+ RT-qPCR positivity while the Chinese sites used RT-qPCR. Therefore, this sensitivity difference is likely due to patients at the Chinese sites who, despite being PCR-positive, express little or no antibody. Overall, we believe that LFA tests will be most useful for surveying antibody levels in patients who are at least 14 days after the onset of symptoms.

Test specificity was found to be excellent across all cohorts. Among the false positive results that were observed, most were found in the IgM channel (12/1438), while only one (1/1438) was found in the IgG channel. This trend may be due to avidity effects arising from the pentavalent nature of IgM. Avidity effects amplify low affinity interactions through multivalency; IgM antibodies that recognize non-SARS-CoV-2 pathogens, but which still share weak structural homology to SARS-CoV-2 antigens present in the test, may still present as test positives. We therefore suggest that patients not rely solely on IgM positivity as a readout of antibody response.

A recent study from UCSF researchers compared several diagnostic testing strategies for SARS-CoV-2 IgG and IgM antibodies, including LFAs originating from manufacturers around the world [5]. These assays were evaluated for various infection time windows (1-5, 6–10, 11–15, 16–20 and >20 days). The Biohit LFA test evaluated in this study exhibited higher sensitivity values for IgG/IgM in patients with infection times >11 days compared with very best LFA and ELISA tests evaluated in the UCSF study (94.1%–100% versus 90.9-100%). Furthermore, the specificity of the Biohit test was comparable to the best performing assays evaluated by the UCSF group (97.6–100% versus 99.1–100%). Therefore, our data support that the Biohit test is an excellent candidate for widespread implementation. That being said, our studies do not represent a head-to-head comparison with assays evaluated by UCSF (i.e., we did not perform studies using the same patient samples), so any comparison must be interpreted with caution.

As described above, we employed a visual intensity score (VIS, Figure 5) to quantify visually the intensity of LFA bands. We found that the VIS strongly correlated with ELISA titer (Figure 6), suggesting that LFA band intensity is proportional to serum antibody levels (although LFAs are often said to function simply as “binary” readouts). That being said, VIS was determined by a trained laboratory scientist, and may not be easily recapitulated by untrained lay users. We also developed an automated intensity score (AIS), wherein photographs of tests taken on a smartphone were analyzed using LICOR image studio.

Although the AIS effectively differentiated strong positive from negative results, it performed poorly to differentiate weak positives from negative results (Figure 7). These results suggest that computer automation could lead to faulty test interpretation. Taken together, we recommend that trained scientists, technicians or health-care professionals interpret physical test results, or test photographs, using tele-medicine or remote diagnosis protocols.

## Conclusion

We report that the SARS-CoV-2 LFA studied herein is suitable for detecting IgM and IgG antibodies to SARS-CoV-2 with high levels of sensitivity and specificity. Test performance is maintained against diverse patient populations and matrices, and in the presence of numerous potentially interfering substances. IgG specificity is near-perfect (99.7-100%) over the entire cohort of 1,892 patient samples. This finding provides reassurance that the studied LFA assay will minimize false positive results. Such false positives are perhaps most problematic from a public health standpoint because they would lead individuals to believe that they are immune or exposed to the virus when in fact they are not. While this LFA cannot predict protective immunity, it can identify people who can be serially assessed for resistance to possible subsequent infection on a population basis. Overall, this LFA shows promise for use as a mobile diagnostic tool, and underscores the potential utility of such a simple and cost-effective assay for cataloging epidemiological data like prevalence and past infection status.

## Data Availability

Data is subject to availability at a future date.

## Acknowledgments

This work was funded by Yale University. We acknowledge Feimei Liu for performing ELISA experiments.

## Competing interests

CM is a cofounder and advisor to Virality Diagnostics. AP is an advisor to Virality Diagnostics. DAS is an advisor and shareholder of Virality Diagnostics, Kleo Pharmaceuticals, Kymera Therapeutics and Biohaven Pharmaceuticals.

## Supplemental figures

**Supplemental Figure 1:**
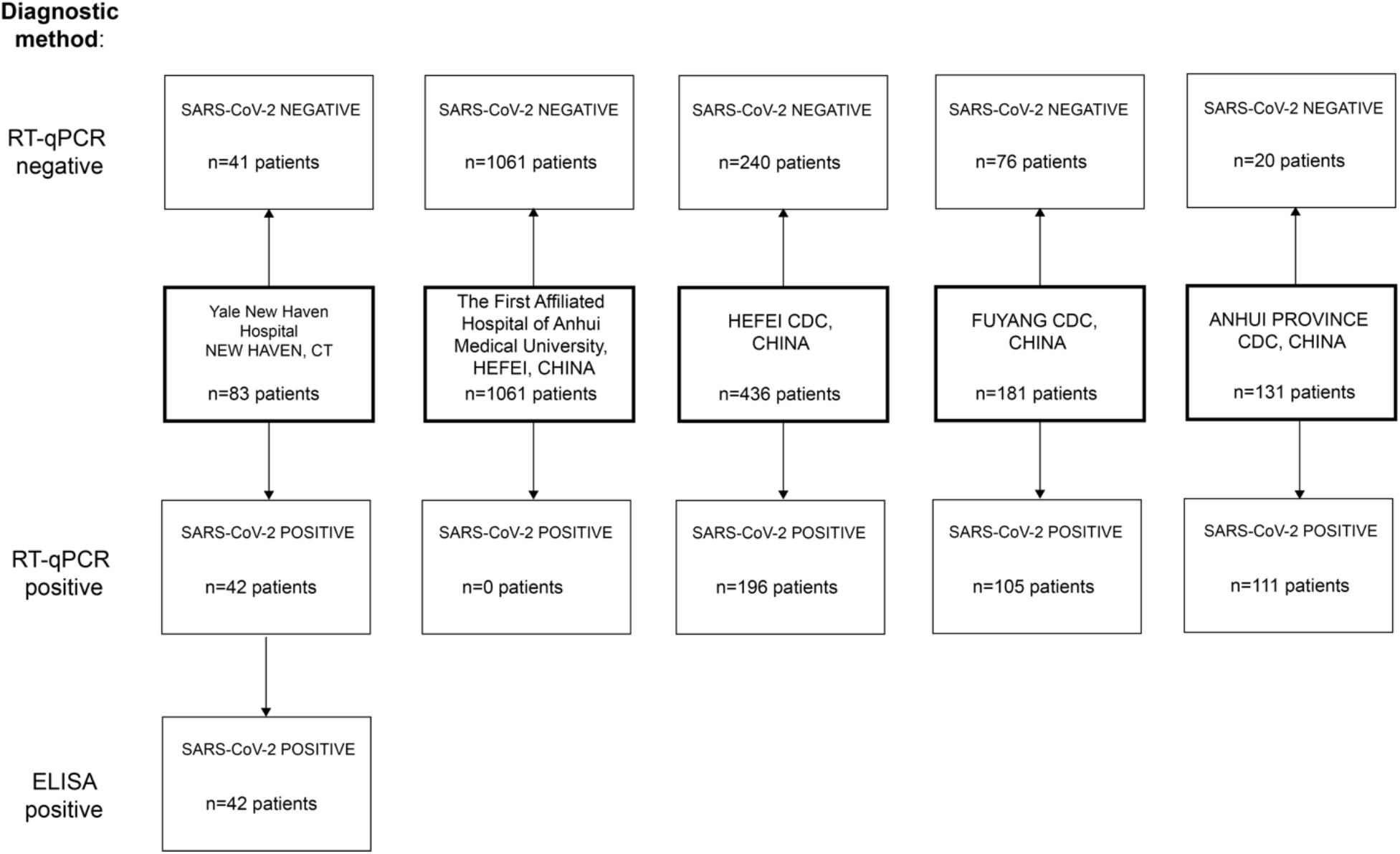
SARS-CoV-2 and control patient selection for study participants.

**Supplemental Figure 2:**
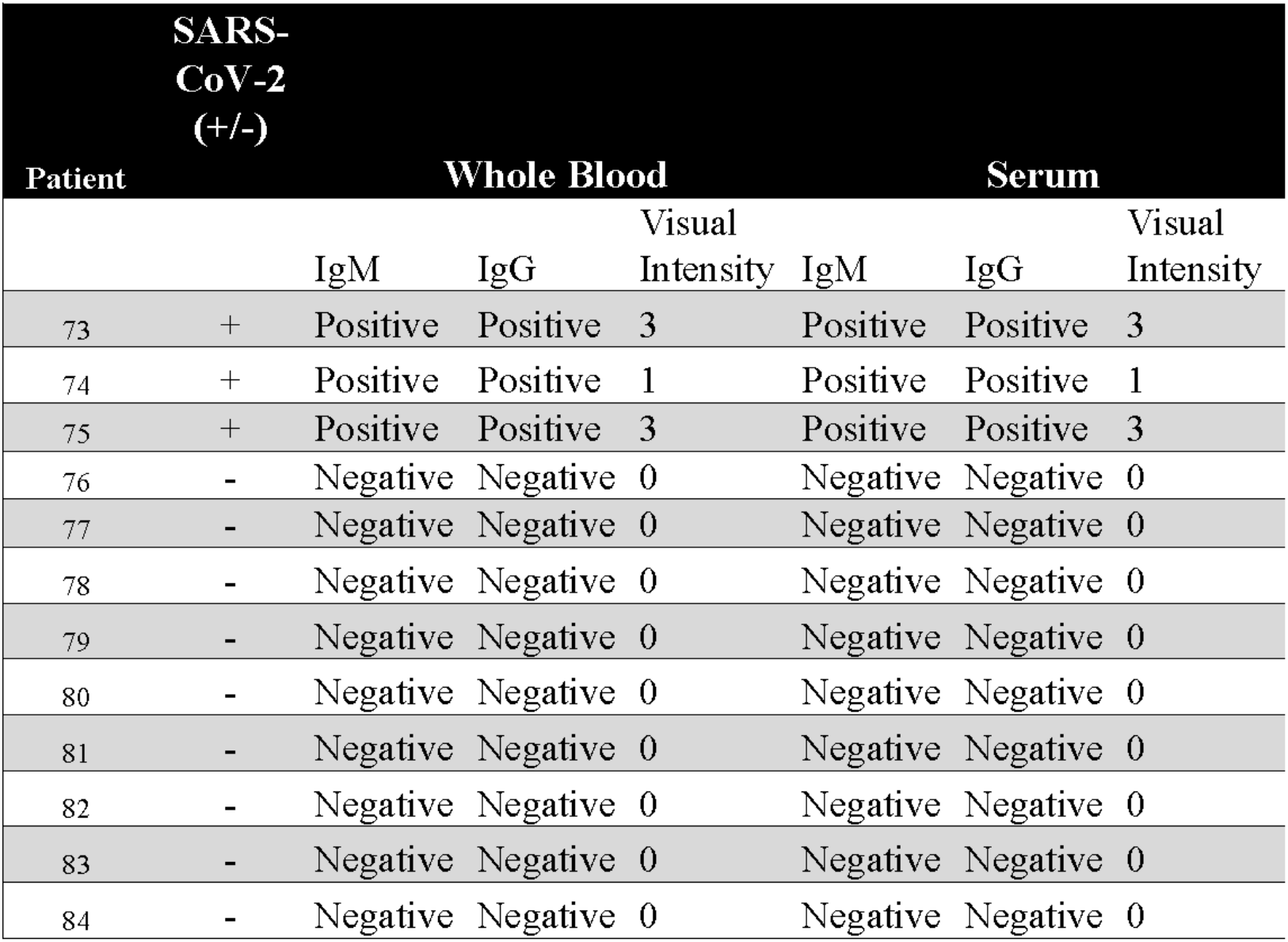
Matrix variability (Whole blood vs. Serum) confirming serum as choice for study matrix

**Supplemental Figure 3:**
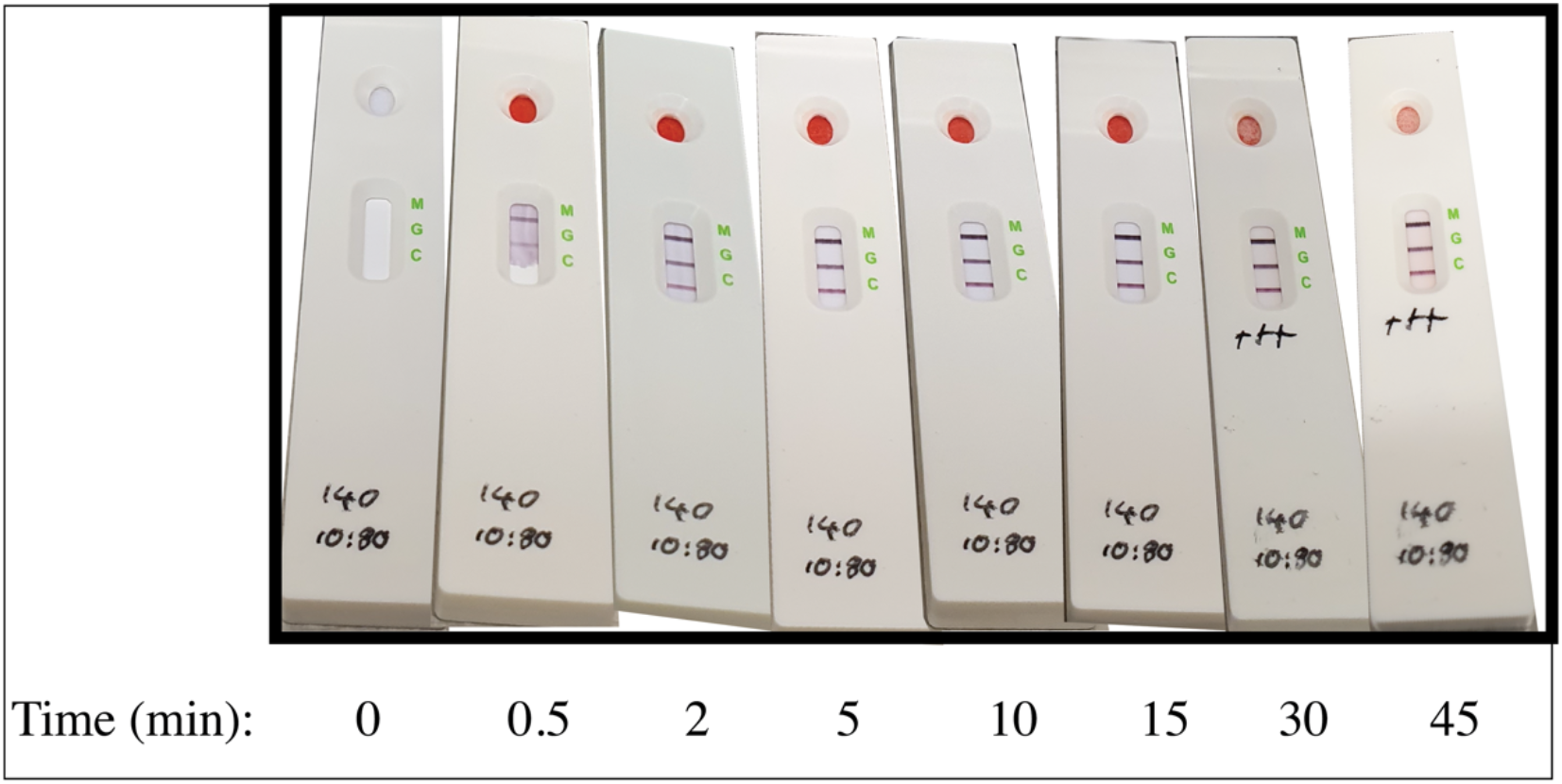
LFA time course evaluation of whole blood samples from SARS-CoV-2 positive patient

**Supplemental Figure 4:**
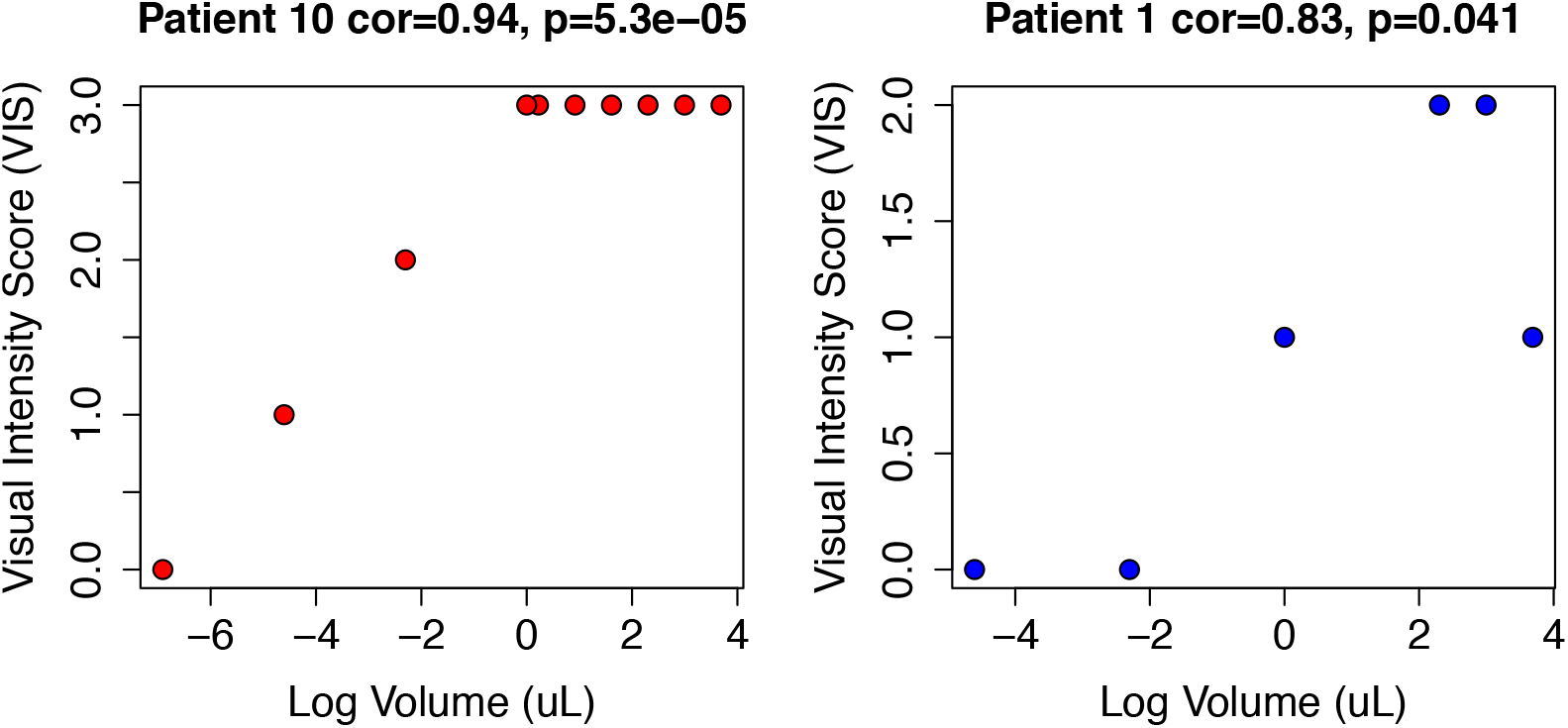
Serum dilution titrations (volume in uL) compared to visual intensity for SARS-CoV-2 patients 10 and 1, confirming 10 uL as the optimal volume for use and suggestion wide variation before alterations in results

**Supplemental Figure 5:**
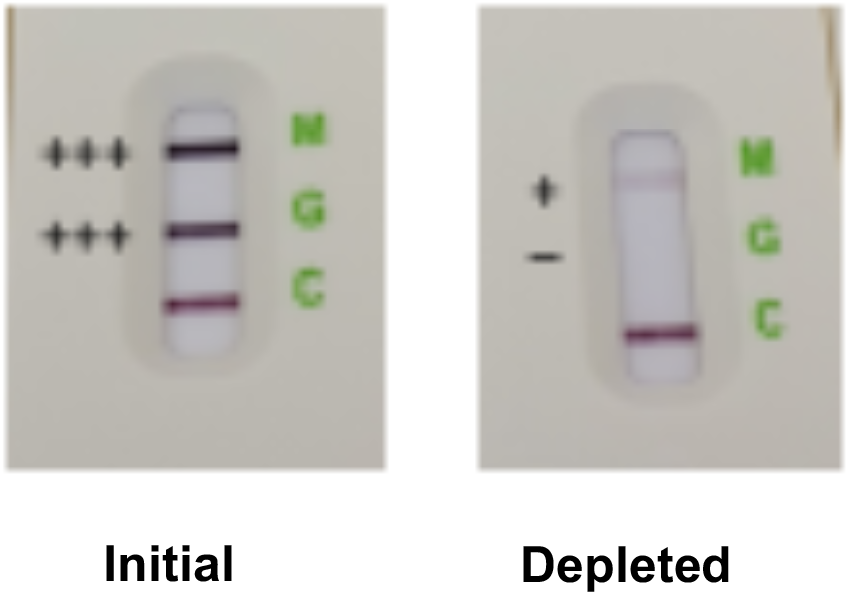
Raw data from IgG depletion studies. Briefly, SARS-CoV-2 positive patient sample was passed over Protein G/A column and subsequent fraction depleted of IgG was tested via LFA.

**Supplemental Figure 6:**
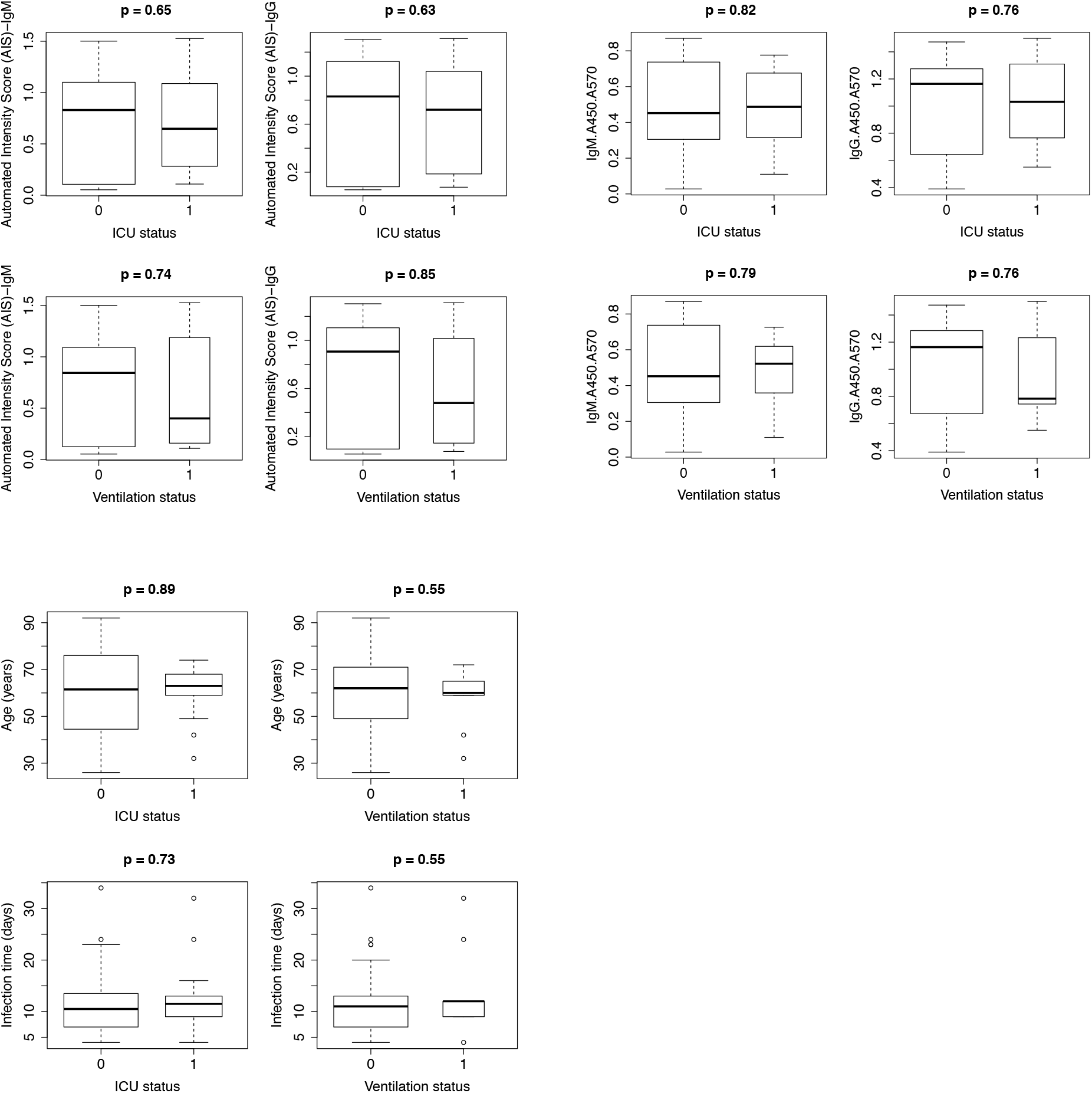
ICU and ventilation status of SARS-CoV-2 patients. 1 = yes, 0 = no.

**Supplemental Figure 7:**
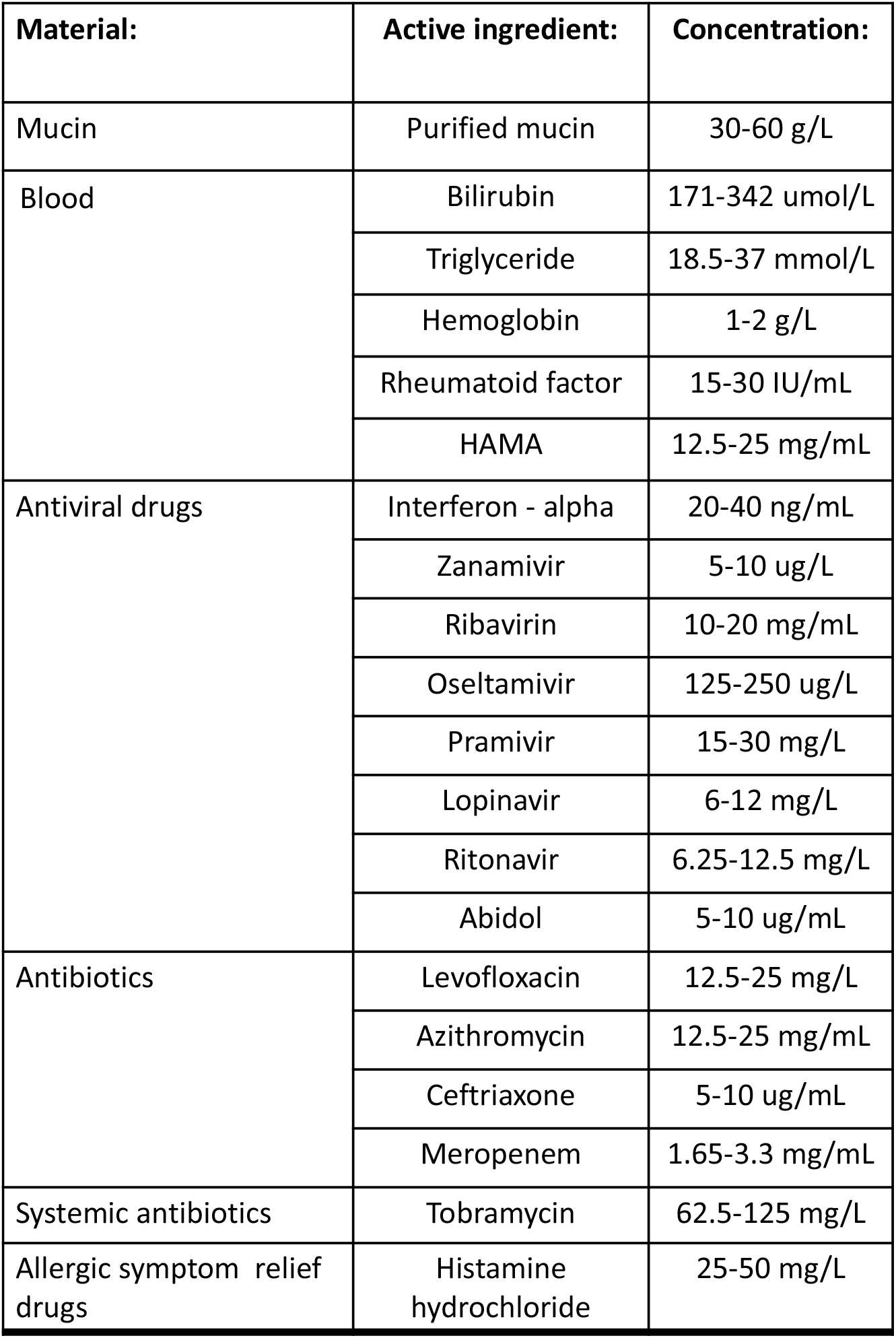
Endogenous interfering substances list for spiking in whole blood, serum and aqueous diluent samples reported no false positives or negatives

## Notes

### Author Declarations

The 83 clinical samples from Yale New Haven Hospital were collected in accordance to the HIC-approved protocol #2000027690. Written informed consent was obtained from all participating patients and healthy controls. Permission was obtained from health and local authorities after they received an explanation of the purpose and procedures of the study. Written informed consent was obtained before participation and blood sampling from all adults and the parents of all participating children less than 18 years. Along with their parents consent, children older than 7 years provided written informed consent. Samples from the Chinese cohorts were obtained from four sites: the First Affiliated Hospital of Anhui Medical University, the Chinese Centers for Disease Control and Prevention in Hefei City (Hefei CDC), Anhui Province (Anhui Province CDC), and Fuyang City (Fuyang CDC). Samples were collected at the First Affiliated Hospital of Anhui Medical University under ethical guidance from the hospital's IRB (Quick-PJ 2020-04-18). Each of the three CDC sites served as a central repository for samples received from local hospitals all of which were collected in accordance with policies outlined by the National Health Commission of the People's Republic of China. Specifically, venous blood samples were collected from any patient displaying symptoms of the novel coronavirus. Those samples were then sent to the governing regional branch of the Chinese Centre for Disease Control and Prevention (CDC), including the three CDC sites participating in this study (Hefei, Fuyang, Anhui). All identifying patient data were removed before data was collected by researchers at Biohit.

